# Lack of latent tuberculosis screening in HIV patients and delay in Anti-Retroviral Therapy initiation in HIV-TB co-infection: A 11-year study in an intermediate TB-burden country

**DOI:** 10.1101/2021.02.15.21251801

**Authors:** Vannesa Yue May Teng, Yan Ting Chua, Eunice En Ni Lai, Shilpa Mukherjee, Jessica Michaels, Chen Seong Wong, Yee Sin Leo, Barnaby Young, Sophia Archuleta, Catherine W.M. Ong

## Abstract

**Objectives:** Tuberculosis (TB) is a common infection in HIV patients. Our study aims to determine the prevalence and characteristics of HIV-TB co-infected patients in Singapore, a high-income, intermediate TB-burden country.

**Methods:** Retrospective data of 11-years was obtained from the National University Hospital (NUH), a quaternary care hospital and the National Centre for Infectious Diseases, the national HIV centre.

**Results:** From December 2005 to December 2016, 48 out of 819 HIV patients and 272 out of 3,196 HIV patients who were managed in NUH and TTSH respectively, were diagnosed with TB. 89.1% (n=285) were males and 2 (0.6%) were screened for latent TB on HIV diagnosis. The median age at TB diagnosis was 47.3 years old (Interquartile range, IQR 41-57). Mean CD4 count at TB diagnosis was 125.0 ± 153.9 cells/mm^3^. 124 (38.6%) patients had CD4 < 50 cells/mm^3^. 41.3% (n=132) of patients had HIV diagnosed at least 6 weeks before TB diagnosis, indicating an opportunity to initiate latent TB preventive therapy. 55.0% (n=176) had HIV and TB concomitantly diagnosed within 6 weeks whilst 2.25% (n=7) had TB diagnosed before HIV. Of those HIV-TB co-infected patients with CD4 ≤ 50 cells/mm^3^, 18 (14.2%) had anti-retroviral therapy (ART) started <2 weeks. TB-related mortality was 5.3% (n=17) and 3.75% (n=12) were lost to follow-up.

**Conclusion:** There is a lack of latent TB screening in HIV patients and a delay in initiation of ART in HIV-TB patients with low CD4 counts in our study. Clinical practices can be further improved for the benefit of outcomes in HIV-TB patients.

## Introduction

In 2019, there was an estimated 10 million new cases of tuberculosis (TB) diagnosed globally, of whom 815,000 of them were among people living with HIV (1). People living with HIV are more susceptible to active TB infection either from reactivation of latent infection or primary infection. HIV infection is estimated to increase the risks of developing TB by 16-27 times compared to those without HIV infection (2). TB remains the leading cause of death in the HIV population worldwide even though TB is both preventable and treatable. Globally, approximately 208,000 people with HIV died from TB in 2019, which may be worsened by the COVID-19 pandemic (1). Reasons for poorer clinical outcomes for HIV-TB co-infected patients include delays in initiation of anti-retroviral therapy (ART) and/or TB treatment, late detection of HIV-associated TB and multi-drug resistance TB (1).

World Health Organization (WHO) promotes preventive action through early systematic screening and treatment for active and latent TB at the time of HIV diagnosis (1). Treatment of latent TB infection (LTBI) is critical to reduce the risk of progression to active TB with an efficacy ranging 60-90% (3). The guidelines currently recommend testing LTBI using either Interferon Gamma Release Assay (IGRA) or Tuberculin skin test (TST) in high-income and upper middle-income countries with estimated TB incidence less than 100 per 100 000 as they are most likely to benefit due to resource availability and TB epidemiology (3). Despite the end TB strategy by WHO that aims to reduce the incidence rate of TB by 90% by 2035 (4), many countries are still not adhering to current guidelines. According to WHO, the percentage of newly diagnosed HIV patients who received preventive LTBI treatment in 2017 in Singapore, Indonesia, India, Philipines, Vietnam, Myanmar, Cambodia was <1%, 16%, 10%, 57%, 31%, 17% and 21% respectively (5).

Singapore is a high-income economy country with an intermediate TB burden (6), and a healthcare system that is consistently ranked as accessible, equitable and cost-effective by international surveys (7)(8). In Singapore, HIV is managed mainly by the National Centre for Infectious Disease (NCID) and is the national HIV referral center. The HIV programme at the National University Hospital (NUH), a quaternary care centre, was established in 2010. In 2018, out of the 1,421 TB cases who had a HIV test performed, 28 cases (2.0%) were found to have HIV-TB co-infection (9). This study was performed to determine the number of active TB cases in HIV patients, and determine the clinical characteristics of this cohort of patients.

## Methods

### Study population

Clinical data were retrospectively obtained from Communicable Disease Centre at TTSH (Tan Tock Seng Hospital), predecessor of the NCID (2005 - 2016) and NUH (2010 - 2016). We included all HIV-positive patients, confirmed by Western blot, of at least age 18, who were treated for TB in both hospitals between December 2005 and December 2016. Data collected were patient demographics including gender, age, comorbidities, HIV diagnosis (date of diagnosis, date of initiation of ART, CD4 count at HIV and TB diagnosis, HIV viral load at HIV and TB diagnosis, screening for LTBI at HIV diagnosis), TB characteristics (date of diagnosis, symptoms and duration of presentation, site of infection, method of diagnosis, presence of cavitation, chest radiograph (CXR) findings, physical examination findings), treatment of TB (TB regime and administration, development of IRIS) and clinical outcome (mortality, lost to follow up, TB recurrence, cause of death).

### Definitions

TB was microbiologically diagnosed by positive *Mycobacterium tuberculosis* cultures or nucleic acid tests. Patients were considered to have clinical TB if they were on TB treatment despite having negative cultures or nucleic acid test. Acid Fast Bacilli (AFB) smear was graded from 0-4, HIV diagnosis was confirmed by Western blotting. CXR were scored 0-10 based on the number of segments involved as previously described by Lawson et al (10). For patients who received treatment in NUH, CXR was scored independently by 3 clinical researchers (Teng V.Y.M, Chua Y.T, Ong C.W.M), and a final score was obtained by the average of 3 scores. For patients managed in TTSH, CXR was scored by a senior clinical researcher (Young, B). LTBI was diagnosed using an interferon gamma release assay (IGRA) (Quantiferon-TB Gold plus; Qiagen) or T-spot TB test (Oxford Immunotec) and/or tuberculin skin test using purified protein derivative (PPD) solution with a cut-off of 10mm as per local practice (9). Multidrug resistant TB was defined as *M*.*tuberculosis* resistant to both isoniazid and rifampicin. Patients were considered lost to follow-up if they did not turn up for follow up visits after starting treatment or if they have left the country without completion of treatment.

### Data Analysis

SPSS software (IBM SPSS, Version 24, USA) and Graphpad Prism (Version 7.0) was used to perform descriptive statistical analyses. Correlation analysis was performed using Spearman’s coefficients and a p<0.05 was considered statistically significant.

### Ethics

Ethics approval was granted by NHG Domain Specific Review Board (reference number 2015/00945).

## Results

During the 11-year study period, 4,015 HIV patients received medical treatment at TTSH and NUH hospital. 48 out of 819 HIV patients and 272 out of 3,196 HIV patients were diagnosed with TB, in NUH and TTSH respectively. Out of 320 eligible patients, the majority were male, with a median age of 47.3 years (IQR 41-57) at TB diagnosis. 93.8% of this cohort of patients were microbiologically-proven TB. Smokers comprised 45% of the cohort and a small proportion had diabetes-mellitus (12.5%) (Table 1).

**Table 1:**
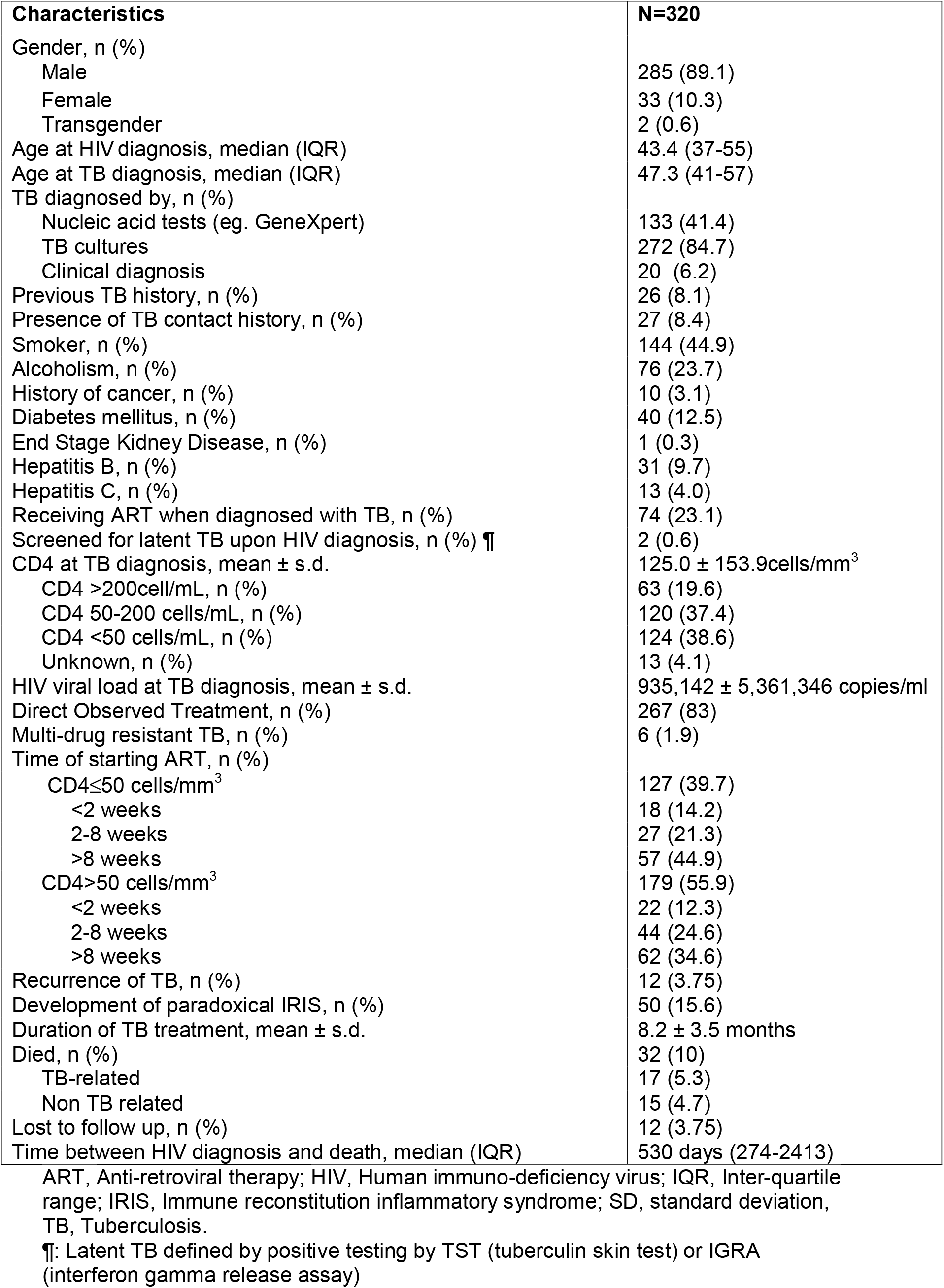
Baseline characterisics of HIV-TB co-infected patients.

| Characteristics | N=320 |
| --- | --- |
| Gender, n (%) |  |
| Male | 285 (89.1) |
| Female | 33 (10.3) |
| Transgender | 2 (0.6) |
| Age at HIV diagnosis, median (IQR) | 43.4 (37-55) |
| Age at TB diagnosis, median (IQR) | 47.3 (41-57) |
| TB diagnosed by, n (%) |  |
| Nucleic acid tests (eg. GeneXpert) | 133 (41.4) |
| TB cultures | 272 (84.7) |
| Clinical diagnosis | 20 (6.2) |
| Previous TB history, n (%) | 26 (8.1) |
| Presence of TB contact history, n (%) | 27 (8.4) |
| Smoker, n (%) | 144 (44.9) |
| Alcoholism, n (%) | 76 (23.7) |
| History of cancer, n (%) | 10 (3.1) |
| Diabetes mellitus, n (%) | 40 (12.5) |
| End Stage Kidney Disease, n (%) | 1 (0.3) |
| Hepatitis B, n (%) | 31 (9.7) |
| Hepatitis C, n (%) | 13 (4.0) |
| Receiving ART when diagnosed with TB, n (%) | 74 (23.1) |
| Screened for latent TB upon HIV diagnosis, n (%) ¶ | 2 (0.6) |
| CD4 at TB diagnosis, mean ± s.d. | 125.0 ± 153.9 cells/mm<sup>3</sup> |
| CD4 >200 cells/mL, n (%) | 63 (19.6) |
| CD4 50-200 cells/mL, n (%) | 120 (37.4) |
| CD4 <50 cells/mL, n (%) | 124 (38.6) |
| Unknown, n (%) | 13 (4.1) |
| HIV viral load at TB diagnosis, mean ± s.d. | 935,142 ± 5,361,346 copies/ml |
| Direct Observed Treatment, n (%) | 267 (83) |
| Multi-drug resistant TB, n (%) | 6 (1.9) |
| Time of starting ART, n (%) |  |
| CD4 ≤ 50 cells/mm<sup>3</sup> | 127 (39.7) |
| <2 weeks | 18 (14.2) |
| 2-8 weeks | 27 (21.3) |
| >8 weeks | 57 (44.9) |
| CD4 >50 cells/mm<sup>3</sup> | 179 (55.9) |
| <2 weeks | 22 (12.3) |
| 2-8 weeks | 44 (24.6) |
| >8 weeks | 62 (34.6) |
| Recurrence of TB, n (%) | 12 (3.75) |
| Development of paradoxical IRIS, n (%) | 50 (15.6) |
| Duration of TB treatment, mean ± s.d. | 8.2 ± 3.5 months |
| Died, n (%) | 32 (10) |
| TB-related | 17 (5.3) |
| Non TB related | 15 (4.7) |
| Lost to follow up, n (%) | 12 (3.75) |
| Time between HIV diagnosis and death, median (IQR) | 530 days (274-2413) |

ART, Anti-retroviral therapy; HIV, Human immuno-deficiency virus; IQR, Inter-quartile range; IRIS, Immune reconstitution inflammatory syndrome; SD, standard deviation, TB, Tuberculosis.
¶: Latent TB defined by positive testing by TST (tuberculin skin test) or IGRA (interferon gamma release assay)

The mean CD4 count at TB diagnosis was 125 ± 153.9 cells/mm^3^ (mean ± S.D.) with 76% of patients having a CD4 count ≤200 cells/mm^3^ at the point of TB diagnosis, indicating profound immunosuppression. Only 2 (0.6%) patients were screened for LTBI upon HIV diagnosis, with equal proportion of unscreened patients above and below CD4 count of 200 cells/mm^3^ (Table 1). Both patients were negative for LTBI by IGRA or TST. Irrespective of CD4 counts, 13%, 23% and 39% of patients had ART started <2 weeks, 2-8 weeks, >2 months of TB diagnosis respectively. Of those with CD4 ≤ 50 cells/mm^3^, only 18 (14.2%) had ART started within 2 weeks.

The most common clinical presentation for TB were fever (75.9%), cough (62%) and weight loss (53.4%). 143 (44.7%) patients had only pulmonary involvement; 39 (12.2%) patients had extrapulmonary TB only and 135 (42.2%) patients had both pulmonary and extrapulmonary TB. In total, 278 (86.9%) eligible patients had pulmonary TB out of 320 patients. The lung was the most common site of TB involvement independent of CD4 count. The most common site of extrapulmonary TB was lymph node involvement for both cohorts above and below CD4 of 200 cells/mm^3^. In patients with CD4 ≤ 200 cells/mm^3^, 87% (n=213) had pulmonary TB whilst 59% (n=144) had extra-pulmonary TB (Figure 1A). However, in those with CD4 > 200 cells/mm^3^, majority comprising 84% (n=53) had lung involvement whilst 39.6% (n=25) had extra-pulmonary TB involvement (Figure 1B).

**Figure 1:**
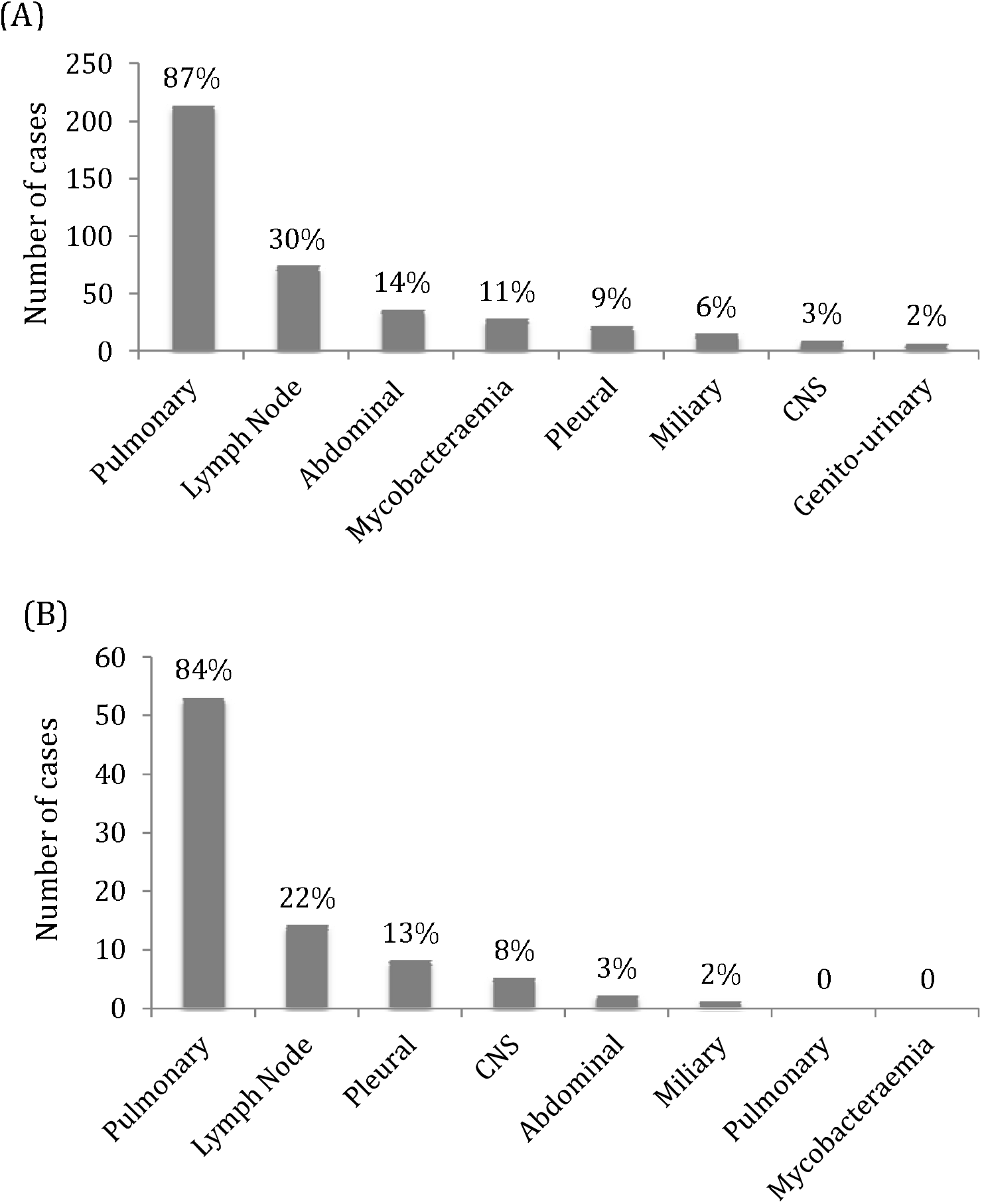
Sites of TB infection in HIV-TB patients. (A) Patients with CD4≤200 cells/mm^3^ at TB diagnosis. (B) Patients with CD4>200 cells/mm^3^ at TB diagnosis

Of those with pulmonary TB, the median CXR score was 4. We found that a total of 84 (30.2%) patients had clear CXR despite the presence of pulmonary TB, most of whom were patients with lower CD4 counts. For patients with CD4 ≤ 200 cells/mm^3^ with lung involvement, CXR can be normal which was seen in 37.9% (Figure 2A), and the same is observed in the relative immunocompetent group (Figure 2B). Most of patients in the CD4 ≤ 200 cells/mm^3^ had positive AFB smear results (Figure 2C) with 10% (n=21) of patients with diffuse lung involvement, having a CXR score of 10 (Figure 2A). However, in patients with CD4 > 200 cells/mm^3^ with lung involvement, only 17% (n=9) patients had clear CXR with a range of AFB smears (Figure 2D), whereas only 3.7% (n=2) patients had diffuse lung involvement (Figure 2B). In both groups, a wide range of AFB smear score from 0 to 4 can be present in patients with a normal CXR (Figure 2E and 2F), indicating that immunocompromised patients can remain highly infectious despite having a normal CXR. Similarly, it was possible to have an AFB smear score of 0 with diffuse TB lung involvement. Interestingly, the AFB smear was positively associated with the degree of CXR score in patients with CD4 > 200 cells/mm^3^ (r = 0.33, p<0.05) but this association was not found in patients whose CD4 ≤ 200 cells/mm^3^ (Figure 2E and 2F).

**Figure 2:**
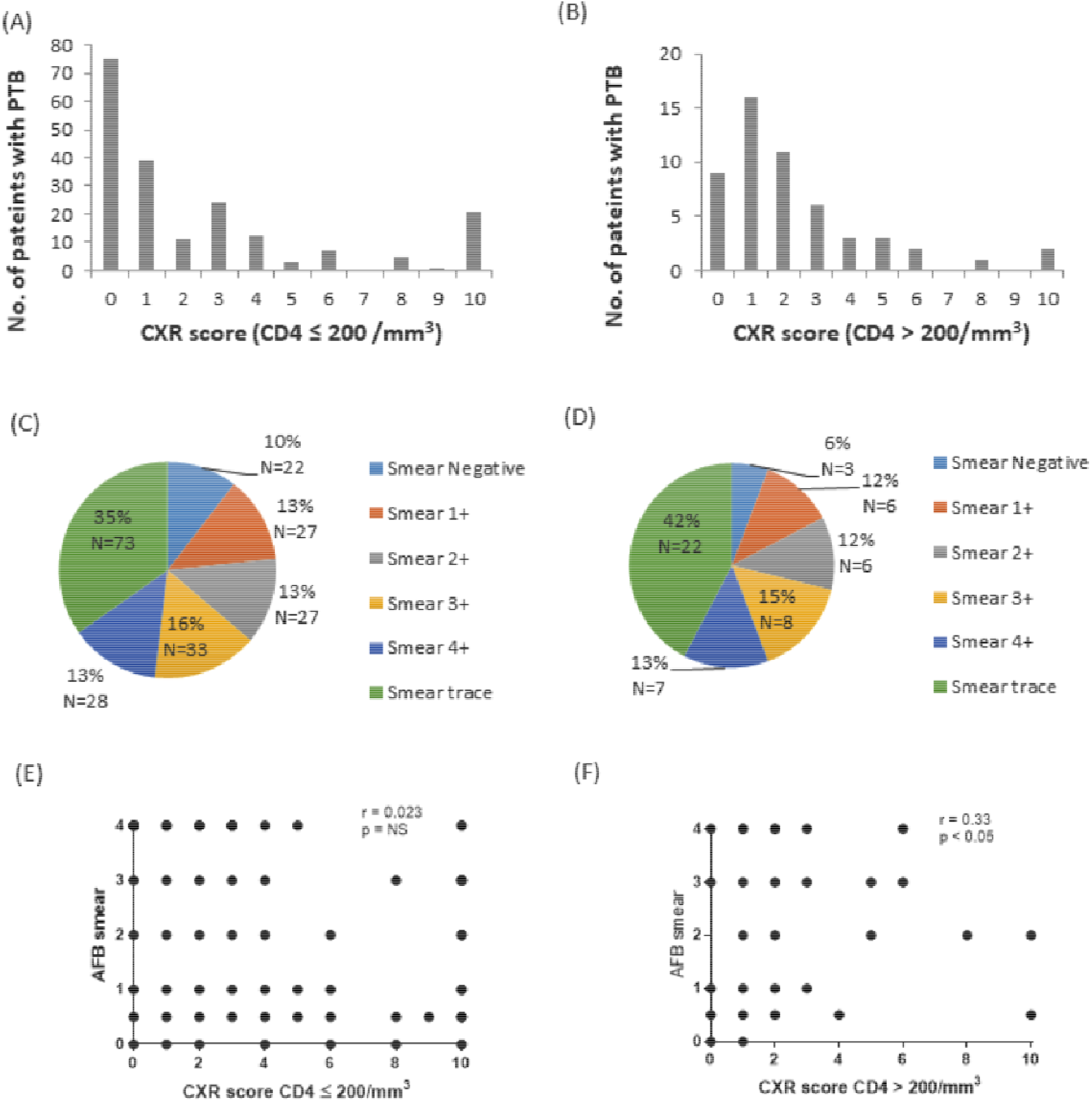
Characteristics of pulmonary TB in HIV patients. (A) Chest radiograph score of patients with CD4≤200 cells/mm^3^ at TB diagnosis. (B) Chest radiograph score of patients with CD4>200 cells/mm^3^ at TB diagnosis. (C) AFB smear quantity in patients with pulmonary TB, with CD4≤200 cells/mm^3^ at TB diagnosis. (D) AFB smear quantity in patients with pulmonary TB, with CD4>200 cells/mm^3^ at TB diagnosis. (E) Association of AFB smear with chest radiograph in patients with CD4≤200 cells/mm^3^ at TB diagnosis. (F) Association of AFB smear with chest radiograph in patients with CD4>200 cells/mm^3^ at TB diagnosis.

41.3% (n=132) of patients had HIV diagnosed at least 6 weeks prior to TB diagnosis with a median time-frame of 46 months (IQR 12-91 months) (Figure 3). The mean duration of active TB treatment was 8.2 ± 3.5 months. The majority of patients (83%) were on directly observed treatment (DOT). A small proportion of patients had TB recurrence (3.8%) and development of immune reconstitution inflammatory syndrome (IRIS) (15.6%). Lost to follow-up rate (3.75%) was low, including those who left the country or defaulted follow-up. All cause-mortality was 10% (n=32), out of whom 53% consist of TB-related mortality (n=17) (Table 1).

**Figure 3:**
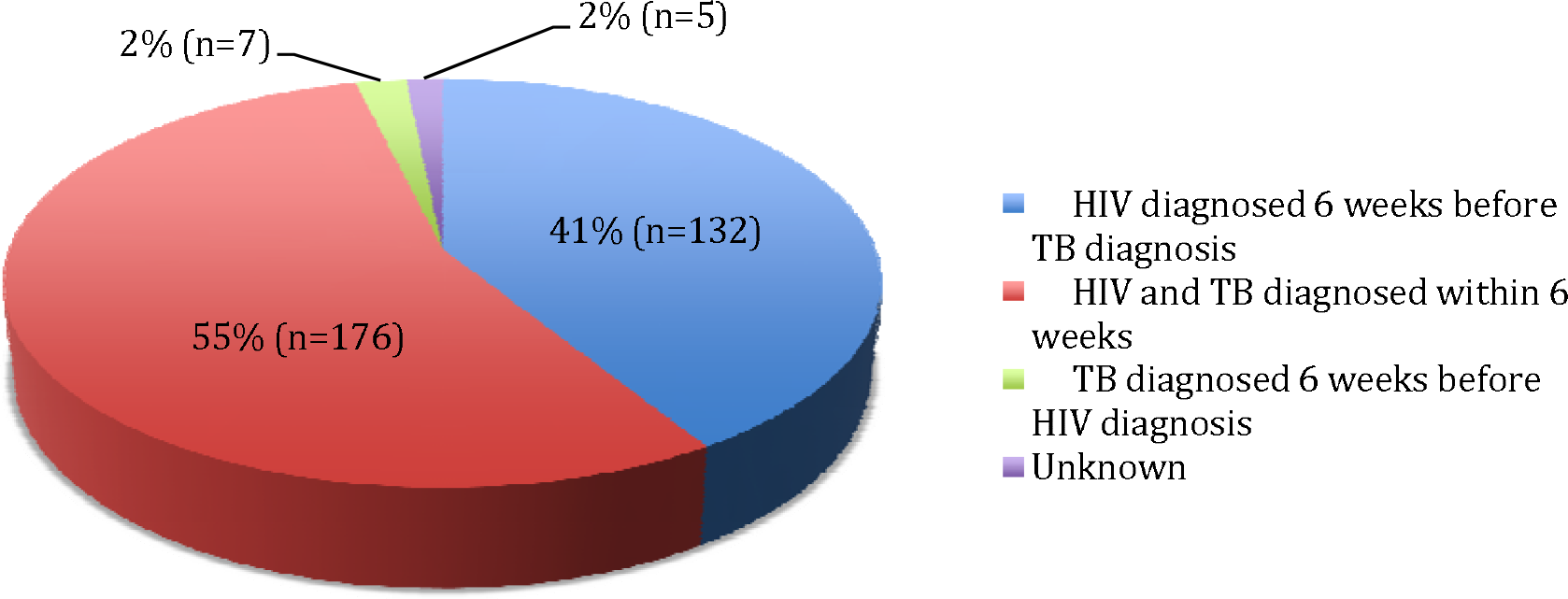
Time to TB diagnosis relative to HIV diagnosis.

## Discussion

In our study, there was a significant proportion of patients (41%) who had HIV diagnosed at least 6 weeks before TB diagnosis. As culture is the gold standard for diagnosis of active TB and culture result takes 2 to 6 weeks to process, our study team considered 6 weeks to be a reasonable time frame to exclude concurrent HIV-TB on presentation (11). Our cohort mainly consisted of clinically advanced HIV disease as 76% had CD4 ≤ 200 cells/mm^3^ at TB diagnosis. However, only 0.6% (n=2) of patients in this study had LTBI screened upon HIV diagnosis, which is a considerably low given WHO recommendations. In developed countries with low TB and HIV burden, the compliance of HIV services in implementing LTBI screening was often inadequate; the LTBI screening rates among HIV patients in United Kingdom, United States and New Zealand was 35%, 32% and 55% respectively (12)(13)(14). The local medical community can further improve on this aspect to prevent active TB in the HIV population.

Our study found higher rates of TB-HIV co-infection in males. This is in keeping with a systematic review done in the European Union which reported that the majority of HIV-TB co-infected cases were male and young adults, aged between 25-45 years (15). We also found that the prevalence of diabetes mellitus (DM) in our cohort was 12% which was similar to a study conducted by Bizune et al in the United States, which reported a prevalence of 9% (16). The high proportion can be explained by diabetes predisposing to TB and ART increasing the risk of metabolic syndrome and thus predisposing to DM (17), and also underscores the increase in DM prevalence in developed countries.

Other significant findings in our study included lower than expected proportion (23%) of patients who were already on ART when diagnosed with TB. This is in contrast with studies in India and Africa which found that 58% and 40% of HIV-infected patient were already receiving ART respectively (18)(19). This difference can be explained by a significant proportion (55%) of our cohort with TB and HIV diagnosed concomitantly within 6 weeks, hence many of them were not expected to be on ART when diagnosed with TB.

Existing studies have reported a variety of chest radiographic appearances of active TB in HIV-positive patients, depending on their level of immunity. In immunocompromised patients, clinical manifestations are atypical as AFB smear can often be positive with normal CXR, or infiltrates are more often present in the lower than the upper lobes (20). 33.5% of our cohort with pulmonary TB had normal CXR which is slightly higher than that reported in other studies which range from 6-21% (21) and may be due to our cohort being more immunocompromised with a mean CD4 count of 125 cells/mm^3^. Therefore, clinicians should exercise caution when using CXR to screen for active TB disease in HIV patients. A high index of suspicion especially in patients presenting with prolonged cough, is required for timely diagnosis of TB in HIV positive patients.

We found 90% of our HIV-TB cohort to be AFB smear positive with 89% in the group with CD4 ≤ 200 cells/mm^3^ and 94% in the CD4 > 200 cells/mm^3^. This is in contrast to previous studies which reported lower prevalence rates (50-70%) with positive AFB sputum smears in HIV patients (21). This difference may be contributed by higher sensitivity used in our laboratory and other studies not reporting trace results from smear tests, leading to lower rates of smear negative results. The mycobacterial burden was also significantly associated with the degree of CXR involvement only in those with CD4 > 200 cells/mm^3^.

WHO recommends initiation of ART for HIV-TB coinfected patients within the first 2 months of starting TB treatment and within 2 weeks in those severely immunocompromised patients with CD4 counts <50 cells/mm^3^. Prompt initiation of ART to HIV-TB co-infected patients reduces mortality especially in advanced HIV disease (22). Our study showed that only 14% of patients with CD4 ≤ 50 cells/mm^3^ had ARV initiated within 2 weeks. In addition, there was a significant proportion (51.7%) of patients who initiated ART more than 2 months of TB diagnosis, out of which 89% were from patients managed prior to 2011. Majority of delay in ART initiation were prior to the revised WHO ART initiation guidelines in 2010 (23). A study by Escada et al in Brazil found that only 5.4% of patients were initiated on ART within 2 weeks of starting TB treatment, however their cohort had a higher median CD4 count of 91 cells/mm^3^ (24).

DOT is recommended by WHO to promote TB treatment adherence as suboptimal adherence has been associated with unfavourable clinical outcomes (25). Our cohort was able to achieve 83% on DOT, with the remaining using self-administered treatment (SAT). There was a wide variability in the success of implementing DOT in other countries, ranging from 24% in South Africa (26) to 91% in Philippines in HIV-TB co-infected patients (27). The main challenges faced worldwide included poor public health infrastructure, poor social support, inadequate funding and lack of awareness about the strategy among practitioners especially in low income, developing countries (27). In our study, TB-related mortality rate was 5.3%. This is much lower than the global rate in 2008 in which TB accounted for about 26% of HIV-related deaths (28). Overall, there is ample room for improvement to comply with international guidelines to reduce mortality in our HIV population.

The main strength of this study is the detailed clinical data collected for analysis over a 11-year period of cohort of 320 patients. Study limitations include missing data given that this was a retrospective study. In addition, although one of our centres is a referral centre for HIV in Singapore, our data may not be representative of the national HIV-TB situation of the country.

## Conclusion

Early diagnosis of LTBI enables prompt effective treatment to prevent TB-associated deaths. Further improvements are needed to offer routine LTBI screening and treatment in all HIV patients, as recommended by the WHO. In our high-income intermediate TB burden country, there remains a good window of opportunity to reduce the prevalence of HIV-TB co-infection, together with prompt initiation of ART to improve treatment outcomes in this population.

## Data Availability

Due to privacy and confidentiality concerns, neither the data nor the source of the data can be made available.

## Acknowledgements

Catherine W. M. Ong is funded by Singapore National Medical Research Council (NMRC/TA/0042/2015, CSAINV17nov014); National University Health System Singapore (NUHS/RO/2017/092/SU/01, CFGFY18P11, NUHSRO/2020/042/RO5+5/ad-hoc/1), iHealthtech in NUS and recipient of the Young Investigator Award, Institut Merieux, Lyon, France.

## Author contributions

Ong C.W.M, Archuleta S. conceived the study. Teng V.Y.M., Chua Y.T., Lai E.E.N., Mukherjee S., Michaels J. and Young B. obtained the data. Teng V.Y.M., Chua Y.T., Lai E.E.N. analysed the data. Teng V.W.M, and Ong C.W.M co-wrote the first draft. All authors edited and approved the final version of the manuscript.

